# Perceived Timing of Cutaneous Vibration and Intracortical Microstimulation of Human Somatosensory Cortex

**DOI:** 10.1101/2021.12.28.21268447

**Authors:** Breanne Christie, Luke E. Osborn, David P. McMullen, Ambarish S. Pawar, Tessy M. Thomas, Sliman J. Bensmaia, Pablo A. Celnik, Matthew S. Fifer, Francesco V. Tenore

## Abstract

**Background:** Electrically stimulating the somatosensory cortex can partially restore the sense of touch. Though this technique bypasses much of the neuroaxis, prior studies with non-human primates have found that conscious detection of touch elicited by intracortical microstimulation (ICMS) lags behind the detection of vibration applied to the skin. These findings may have been influenced by a mismatch in stimulus intensity; typically, vibration is perceived as more intense than ICMS, which can significantly impact temporal perception.

**Objective:** The goal of this study was to evaluate the relative latency at which intensity-matched vibration and ICMS are perceived in a human subject.

**Methods:** A human participant implanted with microelectrode arrays in somatosensory cortex performed a reaction time task and a temporal order judgment (TOJ) task. In the reaction time task, the participant was presented with vibration or ICMS and verbal response times were obtained. In the TOJ task, the participant was sequentially presented with a pair of stimuli – vibration followed by ICMS or vice versa – and reported which stimulus occurred first.

**Results:** When vibration and ICMS were matched in intensity at a “high” stimulus level, the reaction time to vibration was 48 ms faster than ICMS. When both stimuli were perceived as lower in intensity, the difference in reaction time was even larger: vibration was perceived 90 ms before ICMS. However, in the TOJ task, vibratory and ICMS sensations arose at comparable latencies, with points of subjective simultaneity that were not significantly different from zero.

**Conclusions:** Because the perception of ICMS is slower than that of intensity-matched vibration, it may be necessary to stimulate at stronger ICMS intensities (thus decreasing reaction time) when incorporating ICMS sensory feedback into neural prostheses.

## Introduction

Manual touch plays a critical role in object manipulation and performing activities of daily living [1]. One approach to restoring touch to individuals with somatosensory deficits is electrical activation of neurons in the hand representation of somatosensory cortex [2]– [4]. These artificial tactile signals can be used to support and enhance the use of a prosthetic hand [5]. Precise timing of tactile feedback is critical for enabling rapid dexterous object interactions [6]–[8], which raises a key question as to whether intracortical microstimulation (ICMS) can be perceived and/or integrated with a speed resembling that of natural somatosensation. Indeed, lags in ICMS-evoked percepts could impair the utility of sensory feedback [9], [10].

The temporal perception of artificial touch has been studied with peripheral nerve stimulation in humans [11], cortical surface stimulation through electrocorticographic electrodes in humans [12], and ICMS in non-human primates (NHPs) [10], [13]. Despite differences in the mechanisms of neural activation, the temporal processing of sensations evoked by peripheral nerve stimulation was not significantly different than the timing of mechanical stimulation of the skin [11]. On the other hand, even though ICMS bypasses multiple stages of peripheral processing, sensations evoked via stimulation of somatosensory cortex are typically found to be slower to emerge than sensations evoked via mechanical skin stimulation. In NHPs, ICMS latencies were able to match mechanical stimulation latencies only when high currents were delivered through at least four electrodes simultaneously [10]. This inequivalence in latencies could be explained by mismatched intensities between ICMS and mechanical stimulation; intensity has a demonstrated effect on perceptual latency [14]–[19]. Physiological data have demonstrated that stimulus intensity affects the latency and processing rate in sensory pathways [20]; the more intense a stimulus is, the lower the reaction time is.

The objective of the present study was to characterize the latency at which ICMS-evoked sensations are consciously experienced, and to compare the latency of artificial touch to its mechanically-evoked and intensity-matched counterpart in humans. To this end, a human participant implanted with microelectrode arrays in somatosensory cortex performed three sensory tasks: (1) a reaction time task, (2) a temporal order judgment (TOJ) task, and (3) a modality discrimination task. In the reaction time task, the participant was presented with an ICMS pulse train or a cutaneous vibration and provided a verbal response. In the TOJ task, the participant was sequentially presented with a pair of stimuli – vibration followed by an ICMS pulse train or vice versa – and judged which of the two occurred first. In the modality discrimination task, the participant was presented with a stimulus and responded saying if he felt vibration alone, ICMS alone, or both stimuli

## Methods

### Human participant

The following experiments were conducted with a 50-year-old male with C5 (sensory), C6 (motor) ASIA B tetraplegia. He had retained intact somatosensation in his fingertips as indicated by clinical reports. This study was conducted under Investigational Device Exemption (IDE, 170010) by the Food and Drug Administration (FDA) for the purpose of evaluating bilateral sensory and motor capabilities of intracortical microelectrode arrays. The study protocol was approved by the FDA, the Johns Hopkins Institutional Review Board (JH IRB) and the NIWC Human Research Protection Office, and is a registered clinical trial (NCT03161067). The participant gave his written informed consent prior to participation in research-related activities.

### Cutaneous vibration of the hand

A vibratory stimulus (“natural touch”) was delivered on the hand using a miniature electromechanical tactor (type C-2, Engineering Acoustics, Inc.; Casselberry, FL, USA). The tactor was controlled by sinusoidal signals that were digitally generated in MATLAB (MathWorks, Inc.; Natick, MA, USA). Output signals passed through the computer’s audio port and were amplified (Pyle, PTA4 Stereo Power Amplifier). Vibratory inputs were delivered at 300 Hz, which was assumed to primarily activate Pacinian fibers in the skin [21], [22]. We did not anticipate the vibration frequency to impact the results because Pacinian and non-Pacinian afferent fibers have similar diameters and therefore similar conduction velocities [23]. The displacement of the tactor could be varied to change the indentation depth into the skin. The tactor was fastened to the hand using medical tape. The participant reported that he could not hear any noise from the tactor so he did not wear noise-cancelling headphones during experiments.

### Intracortical microstimulation of somatosensory cortex to elicit tactile percepts in the hand

This study involved a human participant who was chronically implanted with microelectrode arrays in the bilateral primary motor cortices and area 1 of the somatosensory cortices (see Fig 1a, and McMullen et al. and Fifer et al. for additional details on these implants [4], [24]). In the present study, only the stimulating electrodes implanted in somatosensory cortex were used. Two of the arrays were implanted in the left somatosensory cortex and one was in the right hemisphere. To access an external stimulator and software, the arrays were wired to skull-fixed transcutaneous metal pedestals. A CereStim R96 (Blackrock Neurotech; Salt Lake City, UT, USA) was used to deliver electrical stimulation to the electrodes. The electrical stimulation waveforms were biphasic, charge-balanced, cathodic-first pulses and were grounded to the metal pedestal. Pulse frequency was set to 100 Hz, total pulse width was 500 μs (200 μs for each phase with a 100 μs interphase delay), and pulse amplitude varied between 30-80 μA. Modulating the pulse frequency of ICMS can also modulate the perceived intensity of ICMS, however the direction of that change varies across electrodes [24], which is why we modulated amplitude instead of frequency. Stimulation parameters stayed within safety limits for minimizing the risk of tissue and/or electrode damage [26]. Stimulation pulse trains were controlled via MATLAB scripts that sent commands to a custom C++-based stimulator interface application.

**Figure 1:**
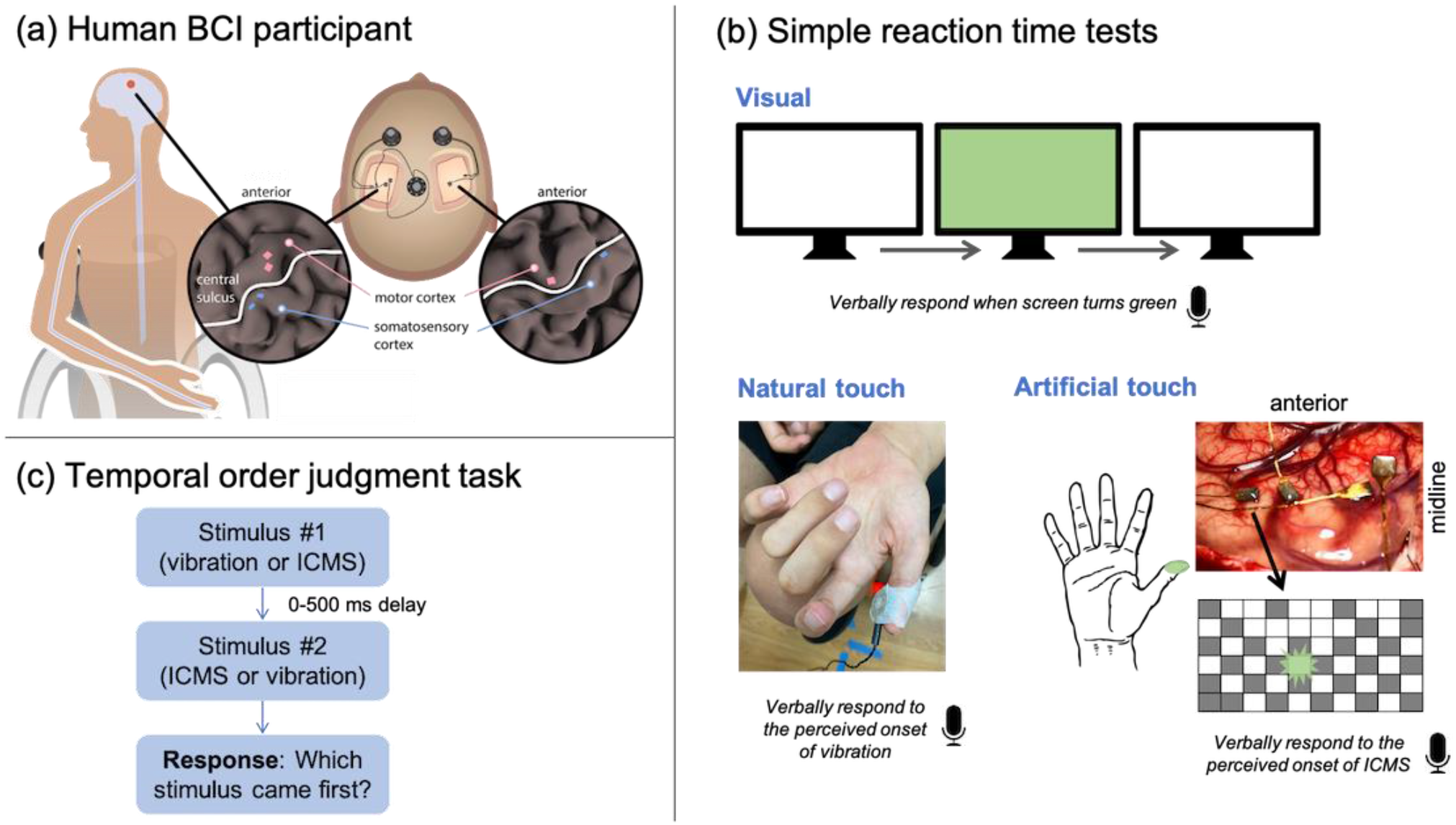
Experimental methods. **(a)** This study involved a human participant with a brain-computer interface. This consisted of two microelectrode arrays in the left hemisphere somatosensory cortex, one array in the right somatosensory cortex, two arrays in the left motor cortex, and one array in the right motor cortex. **(b)** We conducted three versions of a simple reaction time task. During the visual reaction time task, the participant verbally responded into a microphone when he observed a computer screen turning from white to green. During the natural touch reaction time test, the participant verbally responded when he felt a vibratory stimulus on his skin. During the artificial touch reaction time test, the participant verbally responded when he felt tactile percepts elicited by intracortical microstimulation (ICMS). ICMS was delivered via a microelectrode array implanted in the somatosensory cortex. **c)** In the temporal order judgment task, vibration and ICMS were sequentially presented and the participant was asked to respond which stimulus he perceived as occurring first. The stimuli were separated by a stimulus onset asynchrony value between 0-500 ms. We conducted two versions of this task: one in which vibration and ICMS were delivered to the same region of his hand, and one in which vibration and ICMS were spatially offset.

For each site on the hand, ICMS was delivered through a pair of electrodes that had similar projected fields on the participant’s hand. This was done to increase the perceived intensity of ICMS, making it easier and more reliable for the participant to detect. Projected fields were estimated by stimulating through each individual electrode and asking the participant to verbally report the regions of the hand in which he felt a tactile percept. These electrode mappings and the detection thresholds of pairs of electrodes are more thoroughly discussed in Fifer et al [4]. Both electrodes in a pair were always part of the same array, but the electrode pairs were not always the same across all three tasks (Supplemental Figure 1).

### Intensity matching between vibration and ICMS

Because temporal perception is strongly affected by variations in stimulus intensity [14]– [18], [27], we performed perceptual intensity matching between vibration and ICMS on each hand site at the start of each experimental session for all three experimental tasks. Two above-threshold ICMS intensities (one lower, one higher) were chosen for the reaction time task. The lower ICMS amplitude value was generally 1-4.5 dB above the last measured detection threshold for the chosen electrode pair, typically 30 or 45 μA. The higher ICMS amplitude was always chosen to be 80 μA, which was generally 6-9.4 dB above the last measured detection threshold for the chosen electrode pair. An amplitude of 80 μA was used for the TOJ and modality discrimination tasks.

Vibration intensity was matched to a given ICMS intensity using a modified adaptive 2-down, 1-up staircase paradigm with a two-alternative forced choice (2-AFC) presentation of vibration and ICMS in which the participant chose which stimulus was perceived as being more intense. To enable faster identification of perceived matched magnitudes, we modified the adaptive procedure to allow the participant to report if the perceived intensities were the same on any given trial. When the participant perceived both vibration and ICMS to be of the same magnitude, the adaptive staircase paradigm was stopped. Vibration intensity was modulated by changing the indentation amplitude with a constant step size, which translated to approximately 0.8-1.6 dB of the vibration detection threshold. Each location had a slightly different vibration detection threshold, so the amplitude step size was chosen accordingly.

Vibration and ICMS levels were converted to dB using the following relation,

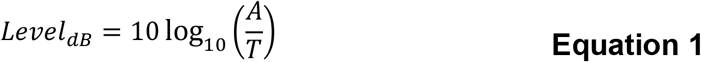

in which *A* is the stimulus amplitude level and *T* is the stimulation detection threshold for the respective stimulation modality at that location. This conversion was done so that the reported stimulation levels could be more easily compared to each other by directly relating them to the detection threshold for each stimulation modality type (i.e., vibration or ICMS).

### Reaction time task

The participant performed a reaction time test for vibratory stimuli, ICMS pulse trains, and visual stimuli (Figure 1b). The participant was instructed to verbally respond with the word “now” into a microphone when he felt a tactile stimulus or observed the visual stimulus (which served as a control condition). Verbal responses were captured using a microphone connected to an H6 audio recorder (Zoom Corporation; Hauppauge, NY, USA) with the raw audio outputted directly to the analog input on a Neuroport Neural Signal Processor (Blackrock Neurotech). The audio waveform was recorded at 30 kHz by the Neuroport, which was also recording the stimulation waveform at 2 kHz. The stimulation and audio waveforms were recorded on the same Neuroport and were aligned using global timestamps. The audio signal was down-sampled to 2 kHz to match the stimulation sampling rate. Reaction time was measured as the time difference between stimulation onset and verbal response onset. Verbal response onset was defined as when the recorded audio signal exceeded a manually-defined threshold, which was approximately six standard deviations above baseline.

Each stimulus lasted for 500 ms with a random delay of 3-7 s between each trial. Reaction times that fell below 100 ms or above 1000 ms were considered physiologically implausible and excluded from the analysis [28], [29]. There were 54 trials with responses outside of this range (18 vibration, 17 ICMS, and 19 visual). Experiments were performed in blocks of 10 trials to minimize the occurrence of perceptual adaptation [30]. After removing outlier responses, a total of 287 trials were collected for visual stimuli, 282 trials were collected for vibration, and 279 trials were collected for ICMS. Data were collected across eight sessions, approximately once a week, over the course of three months.

Only one stimulus type (vibration, ICMS, or vision) was presented at a time. During the visual reaction time test, the participant viewed a monitor and responded when the screen changed colors. For the vibration and ICMS tests, we tested five different sites across both hands, at two different intensity levels per site (Supplemental Table 1). The participant focused his gaze on his hand while receiving vibration or ICMS.

### Temporal order judgment task

To evaluate the temporal synchrony of cutaneous vibration and ICMS, we conducted a two-alternative forced choice TOJ task (Figure 1c). In this task, vibration and ICMS were sequentially delivered and the participant was asked which stimulus he perceived as occurring first. The stimuli were separated by a stimulus onset asynchrony (SOA) value of 0, ±50, ±100, ±200, ±300, or ±500 ms. Positive SOA values indicate that ICMS occurred before vibration, and negative values indicate that vibration occurred first. Any hardware latencies were accounted for such that, if commanded to be delivered simultaneously, the onset of vibration and ICMS were aligned within 1 ms. The start of each trial was indicated by a brief audio cue, and each stimulus lasted for 200 ms. We chose three sites on the hands to apply vibration and subsequently picked stimulating electrodes that elicited ICMS percepts in the same region or spatially offset regions of the hand, depending on the experimental condition (Supplemental Table 1). The order of application of the 11 SOA values was randomized and each SOA was tested 20 times per site. In one instance, in which vibration and ICMS were delivered to the right thumb, we tested each SOA 36 times; when we fit the logistic curve to the raw experimental data after just 20 repetitions, the R2 was only 0.63, and we anticipated that additional trials would smooth out any initial noise in the raw data. Each SOA was tested one time per site per experimental block; blocks were kept short to minimize perceptual adaptation. Data were collected in 18 sessions over the course of eight months.

To compare the TOJ results across conditions, we used a parametric bootstrap method with 1000 replications [31]–[33]. This involved generating a synthetic dataset by sampling from a binomial distribution *B*(*n, p*), in which *n* = 20 trials for each SOA, and *p* = the probability that the participant reported that ICMS came first based on a sigmoidal curve (Equation 2) fit to the raw experimental data. Sigmoidal curves were then fit to each synthetic data set to extract two outcome measures: the point of subjective simultaneity (PSS) and just noticeable difference (JND) (Supplemental Figure 2a). The PSS equaled the SOA that corresponded to when the participant reported that he felt ICMS first in 50% of trials. In other words, the PSS represented the temporal offset that resulted in vibration and ICMS being perceived as maximally simultaneous. The JND was equal to the difference in SOA between the 25% and 75% points divided by two [34].

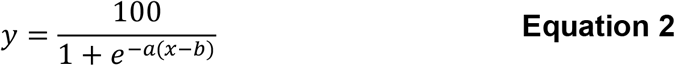

### Modality discrimination task

To further interpret the reaction time and TOJ results, we performed an additional test to verify that vibratory stimuli and ICMS were clearly perceived and did not interfere with one another. In this test, we delivered either a combination of vibration and ICMS in the same region of the hand (“multiplexed”), vibration alone, ICMS alone, or no stimulus. The participant’s task was to report which class(es) of stimulus was presented. If both stimuli were presented, he did not have to specify which came first. Each stimulus lasted 200 ms. To assess interference effects, i.e. to evaluate if it was more difficult to detect two stimuli that were simultaneous vs. temporally offset, we varied the SOA during trials when both vibration and ICMS were delivered; the stimuli were simultaneous, vibration preceded ICMS by 200 ms, or ICMS preceded vibration by 200 ms. This temporal offset of 200 ms was selected because it ensured that one stimulus ended before the next began. Each trial began with an auditory cue followed by a one second delay and the stimuli. ICMS amplitude was always 80 μA and the vibration amplitude was matched in perceived intensity using the approach described above. This task was performed at four locations across both hands (Supplemental Table 1), each in different experimental blocks. Each experimental block consisted of 20 trials (five repetitions of each condition). Data were collected over four sessions, approximately once every two weeks over the course of two months. There were a total of 30 trials per condition on the right thumb and right index finger and 15 trials per condition at the other two locations.

### Statistical analyses

We performed two-sample two-tailed Wilcoxon rank-sum tests to compare the reaction times of the tactile stimuli (i.e., vibration vs. ICMS) as well as tactile stimuli vs. visual stimuli. To compare low-vs. high-intensity stimuli, we ran paired-sample one-tailed Wilcoxon signed-rank tests, with the hypothesis that more intense stimuli would be perceived more quickly. Data normality was not tested because we used nonparametric statistical tests.

For the TOJ results, we fit a probability density function to the 1000 bootstrapped PSS values and calculated the proportion of samples that fell above zero, which was assumed to be the one-tailed p-value (Supplemental Figure 2b). A two-tailed test, to evaluate whether the PSS was significantly above or below zero, was performed by doubling the one-tailed p-value. The PSS data did not deviate significantly from normality (Shapiro-Wilk test, p>0.05). Additionally, to compare the PSS and JND values between the spatially co-located vs. offset conditions, we calculated the proportion of overlap (taken to be the p-value) between the probability density functions of the two conditions (Supplemental Figure 2c).

To analyze the modality discrimination task results, we ran one-sample binomial tests to see if the participant’s ability to correctly identify a stimulus was better than chance. We set the chance level to the response proportions we observed rather than 25%. Though the participant was asked to give one of four answer choices (no stimuli, ICMS only, vibration only, or multiplexed), there were a different number of trials per each of those conditions due to the temporal offsets that were also tested with multiplexed stimuli. If the participant had chosen his responses purely based on what he had learned about the frequency of each stimulus condition, the chance levels would have been 90/540=0.167 for no stimuli, ICMS only, vibration only, and 270/540=0.50 for all of the multiplexed stimuli. Instead, we found that the proportion of times he reported feeling no stimuli was 0.1704, ICMS only was 0.1815, vibration only was 0.2241, and multiplexed stimuli was 0.4241. The expected vs. observed distributions were statistically significantly different, per a two-tailed Chi-square test (p=0.001). Therefore, we chose to use the participant’s response proportions in the statistical tests as a compromise between 25% (based solely on the four response categories) and 16.7/50% (based solely on the number of trials per condition). The binomial tests were one-tailed because we hypothesized that the participant could discriminate between stimuli at an accuracy above chance.

To compare the modality discrimination accuracy between the simultaneous and temporally offset multiplexed conditions, we ran two-sample binomial proportion tests. The tests were one-tailed because we hypothesized that the participant could discriminate between stimulus types better when the stimuli were temporally offset. Significance levels in all statistical analyses were set to α=0.05 and Bonferroni corrections were applied for each task. MATLAB was used to perform these analyses.

## Results

### Reaction time task

We observed reaction times of 366 ± 106 ms (mean ± standard deviation) for the low-intensity vibrations and 352 ± 111 ms for the high-intensity vibrations (Figure 2 and Supplemental Figure 3). The reaction times were faster for the more intense stimuli (p<0.05), as expected. Reaction times were consistent with previously reported values from studies in which participants responded using a button press, which ranged from 177-400 ms [12], [35], [36], particularly considering that vocal responses tend to be 60-80 ms slower than manual ones [37]. Reaction times to ICMS pulse trains were 456 ± 143 ms for low-intensity stimuli and 400 ± 140 ms for high-intensity stimuli. Similar to prior studies [14]–[19] and our results with vibratory stimuli, the effect of intensity on ICMS reaction time was also statistically significant (p<0.001). Importantly, reaction times to ICMS were slower than reaction times to vibration (p<0.001 at both low and high intensities).

**Figure 2:**
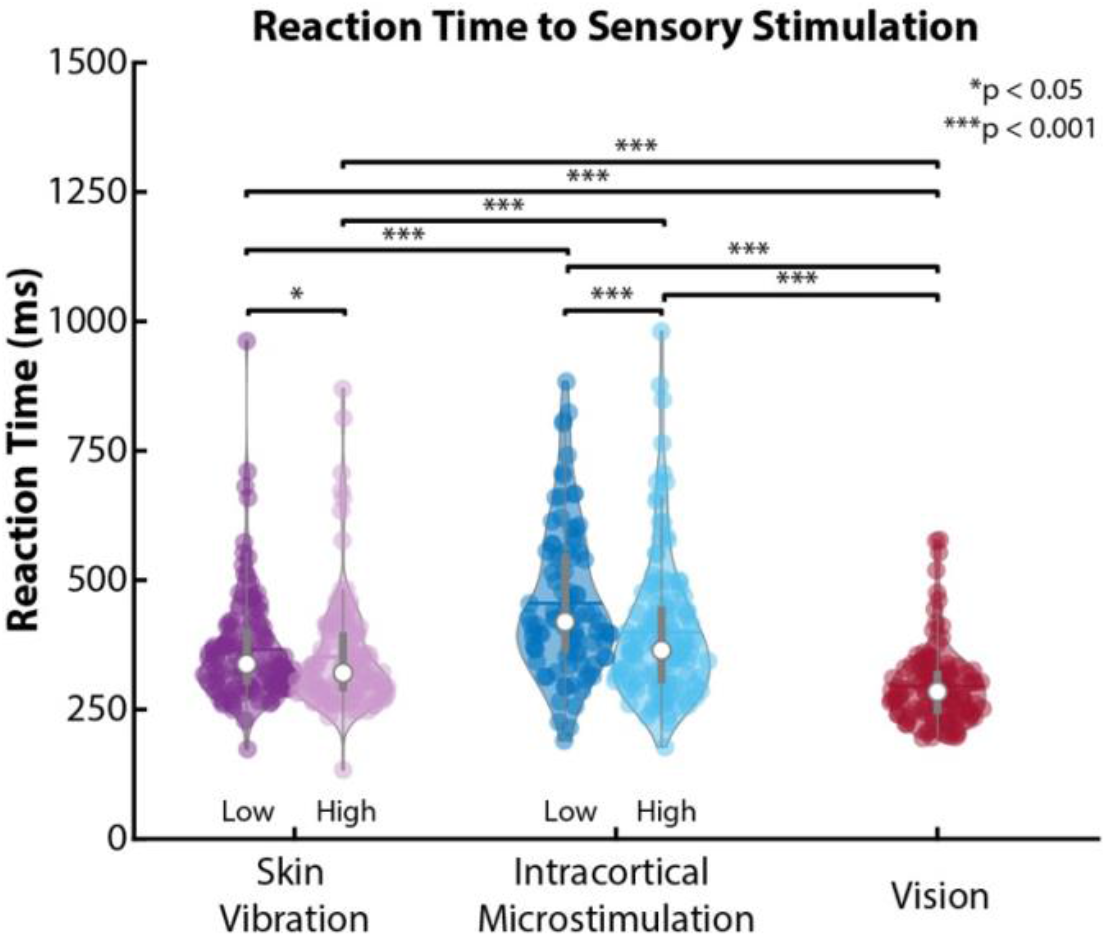
Reaction time test results for vibratory stimuli, intracortical microstimulation (ICMS), and visual stimuli. Each point on the graph represents one trial and the asterisks indicate statistically significant differences. “Low” and “high” represent the two intensity levels tested for both vibration and ICMS (both levels being higher than threshold amplitudes). The vibration and ICMS results are collapsed across the five different test sites.

To further benchmark the participant’s reaction times with respect to individuals without sensorimotor deficits, we measured his reaction times to visual stimuli. We found that the mean reaction time of 295 ± 75 ms was comparable to other individuals who were normally-sighted (around 330 ms [37]). The participant’s reaction time to visual stimuli was significantly faster than his reaction to tactile stimuli (p<0.001 at either intensity level for either tactile stimulus).

### TOJ task

When vibration and ICMS elicited percepts in spatially offset regions of the hand, the PSS was -12 ± 25 ms for one pair of stimulus locations (vibration on right ring, ICMS on right thumb), -57 ± 29 ms for another (right thumb, right ring), and -50 ± 26 ms for a third (left thumb, left index) (Figure 3a and Supplemental Figure 4). The negative values indicate that vibration needed to occur before ICMS for the two stimuli to be perceived as simultaneous. While the PSSs were systematically less 0, none were significantly so, meaning that vibration and ICMS were perceived as maximally simultaneous when presented around the same time.

**Figure 3:**
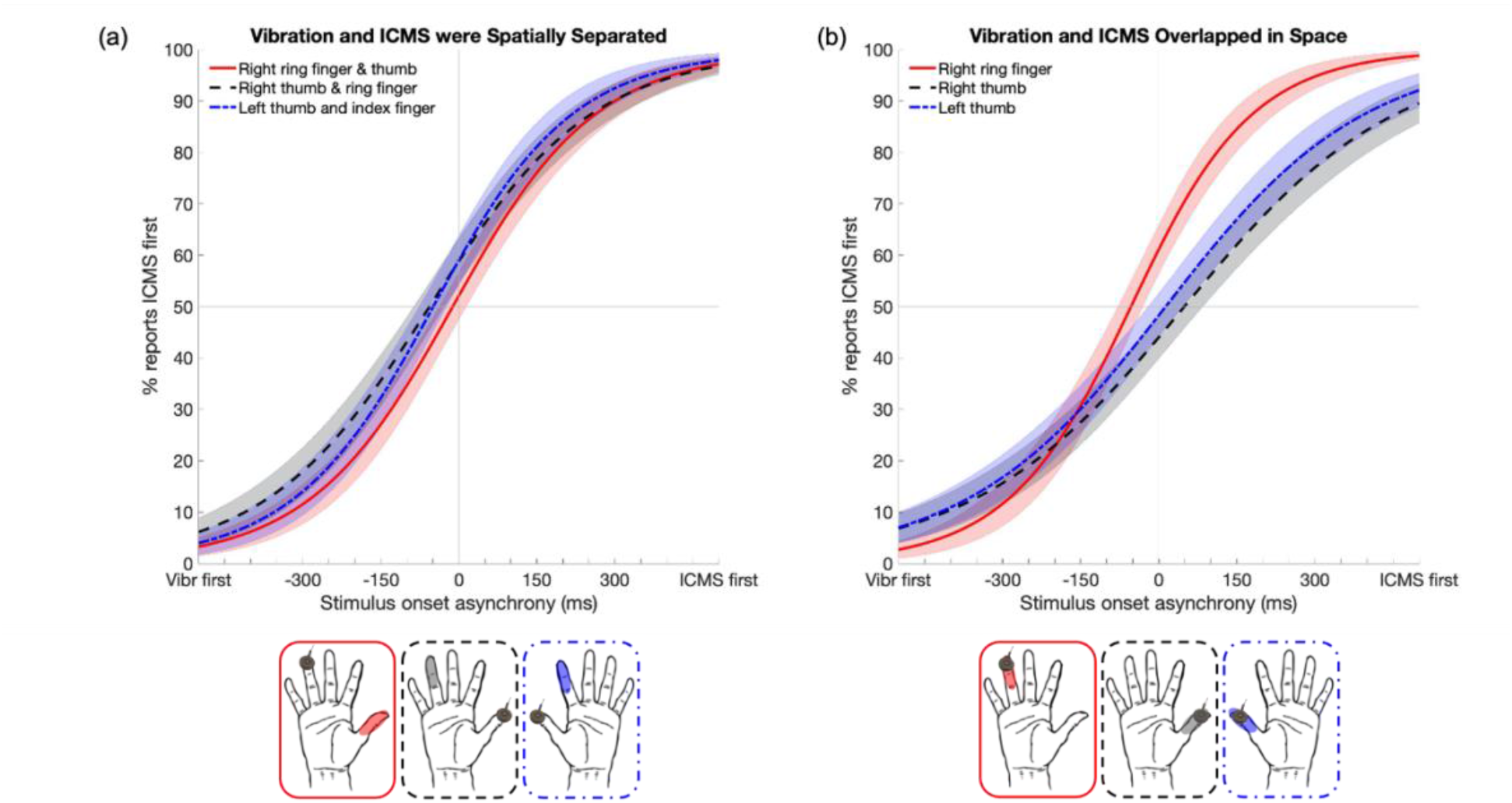
Temporal order judgment task graphical results. Positive stimulus onset asynchrony values indicate that intracortical microstimulation (ICMS) occurred before vibration, and negative values indicate that vibration (‘Vibr’) occurred first. Sigmoidal curves were fit to the raw data and a bootstrap analysis was performed to get the error bars, representing standard deviations, for each curve. In the bottom row, the image of tactor indicates where vibration was delivered and an opaque oval indicates where ICMS was delivered. For all conditions, the point of subjective simultaneity (corresponding to a 50% “ICMS first” reporting percentage) was not significantly different from 0 ms. **(a)** Vibration and ICMS were delivered to different regions of the same hand. Solid red line: vibration of right ring finger, ICMS of right thumb. Dashed black line: vibration of right thumb, ICMS of right ring. Dashed-dotted blue line: vibration of left thumb, ICMS of left index finger. **(b)** Vibration and ICMS were delivered to overlapping regions of the hand. Solid red line: vibration and ICMS were both delivered to the right ring finger. Dashed black line: vibration and ICMS were both delivered to the right thumb. Dashed-dotted blue line: vibration and ICMS were both delivered to the left thumb.

When vibration and ICMS were co-located, PSSs were -54 ± 24 ms for the right ring finger, +50 ± 34 ms for the right thumb, and +13 ± 34 ms for the left thumb (Figure 3b). Again, PSSs were not significantly different from zero after Bonferroni correction. The PSS values in the spatially offset vs. co-located conditions were also not significantly different (p > 0.05).

Finally, the JNDs suggested that the participant was sensitive to changes in timing over a time scale of hundreds of milliseconds. When vibration and ICMS were spatially offset, JNDs were 155 ± 25 ms, 176 ± 28 ms, and 152 ± 27 ms for the three conditions. When vibration and ICMS were co-located, the JNDs were 133 ± 23 ms, 230 ± 40 ms, and 216 ± 37 ms. The JND values in the spatially offset vs. co-located conditions were not significantly different (p > 0.05).

### Modality discrimination task

The participant was able to reliably identify the stimulus modalities delivered across the four sites tested (Figure 4 and Supplemental Figure 5). “Vibration only” conditions were classified correctly 86% of the time and “ICMS only” conditions were correctly classified 93% of the time (chance = 25%). When both stimuli were presented simultaneously (the “multiplexed” condition in Figure 4), the participant’s accuracy dropped to 69% correct, but was still significantly higher than chance (p<0.001). When the participant misclassified trials containing both stimuli, he was most likely (90%) to report only perceiving vibration, despite stimulus levels being matched in perceived intensity. When the stimuli were temporally offset by 200 ms, his classification accuracy significantly improved to 83% when vibration preceded ICMS (p=0.011) and 91% correct when ICMS preceded vibration (p<0.001).

**Figure 4:**
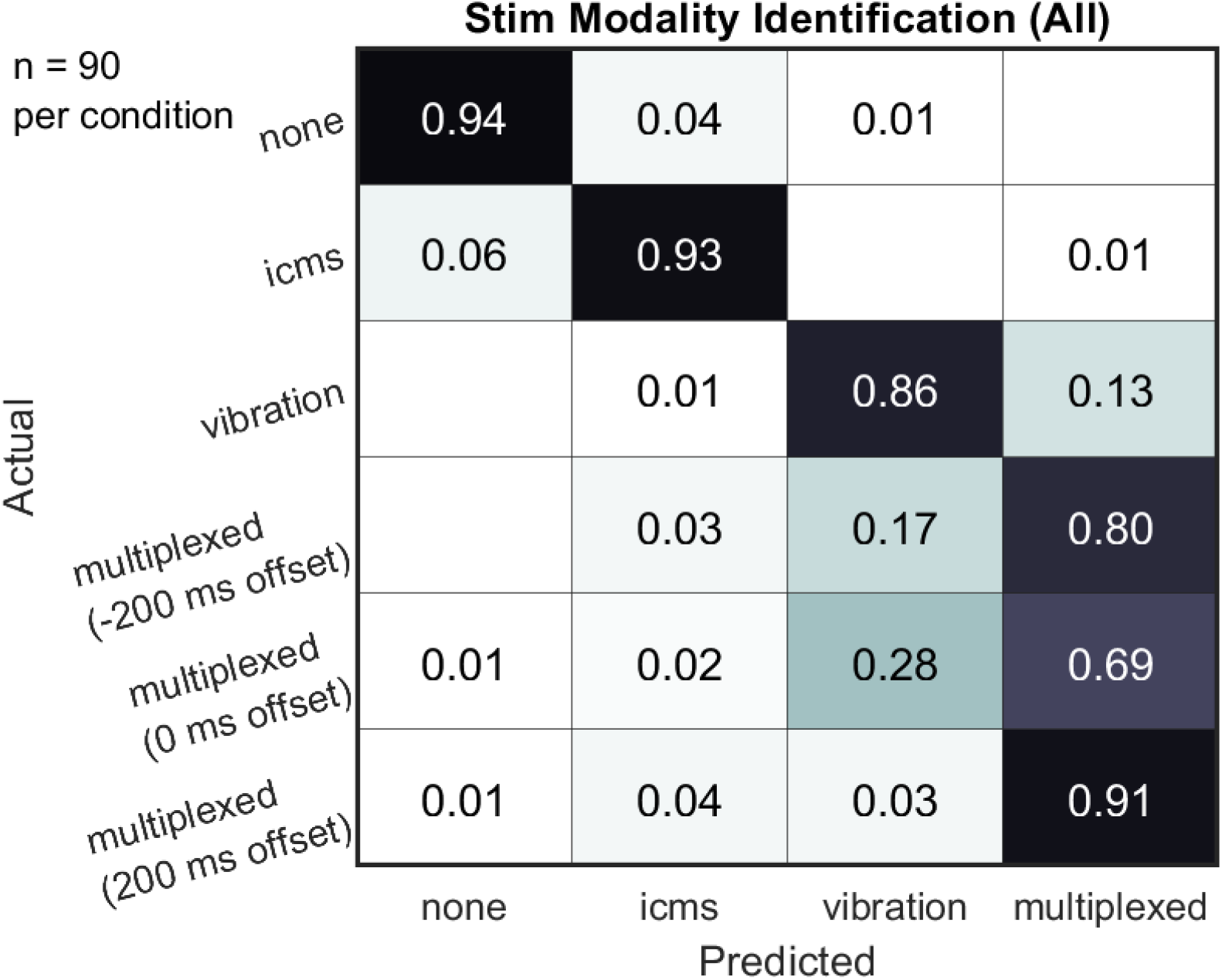
Modality discrimination task results, collapsed across the four different sites on the hand. During the “multiplexed” condition, both vibratory stimuli and ICMS were delivered. A negative temporal offset during the multiplexed condition signifies that vibration preceded ICMS and a positive offset indicates that ICMS preceded vibration. The participant was able to correctly identify trials with only vibration and only ICMS 86% and 93% of the time, respectively. The participant’s ability to accurately identify both stimuli modalities was lower compared to identifying a single stimulus modality, with an accuracy of 69%. When vibration and ICMS were temporally offset by 200 ms, the participant’s classification accuracy significantly improved to 83% when vibration preceded ICMS (p=0.011) and 91% correct when ICMS preceded vibration (p<0.001).

## Discussion

The goal of this study was to evaluate the relative latency at which intensity-matched artificial and natural touch sensations are perceived. To this end, we conducted reaction time tests and TOJ tests with a human participant implanted with stimulating microelectrodes in his somatosensory cortex [4], [24]. Consistent with previous studies [10], [12], [13], we found that the reaction time to high-intensity vibration was 48 ms faster than high-intensity tactile percepts elicited by cortical stimulation, even when the two stimuli were matched in perceived intensity. When both vibration and ICMS were delivered at a lower intensity level, the difference in reaction time was even larger: vibration was perceived 90 ms before ICMS. However, in the TOJ task, vibratory and ICMS sensations seemed to arise at comparable latencies, with estimated PSS values which were not significantly different from zero.

### Vibration is perceived more quickly than ICMS

In this study, we found that the reaction time to vibration was faster than that of ICMS, and. This result refuted our original prediction that the reaction time to artificial touch would be faster than vibration if the stimuli were intensity matched, but is consistent with previous findings [10], [12], [13]. If vibration and ICMS do not need to be matched in perceived intensity, the reaction time to ICMS can be made faster by increasing ICMS current and/or simultaneously stimulating through multiple electrodes [10]. However, the influence of stimulus intensity on reaction time will eventually saturate [38]. Similarly, we found that the reaction time decreased as stimulus intensity increased for both vibration and ICMS. There were also differences in ICMS reaction times across hand sites, again consistent with prior studies [10] and expected because we did not intensity-match ICMS across sites.

Delayed perception of ICMS may be due to the fact that ICMS simultaneously activates somatosensory cortex neurons in a nondiscriminatory and therefore unnatural spatiotemporal manner [13]. In contrast, in a prior study, temporal perception did not vary significantly between touch elicited by peripheral nerve stimulation and mechanical skin indentations [11]. Though peripheral nerve stimulation bypasses mechanoreceptors in the skin, the afferent signals still pass through the spinal cord and thalamus, which may aid in their integration and more rapid interpretation. Additionally, the participant in this study verbally described ICMS sensations as “pressure” or “tingling,” but he reported that it felt different than normal pressure and different than vibration. None of the electrodes in any of the participant’s arrays elicited an ICMS sensation that was described as resembling vibration in prior characterization experiments, even when testing a variety of stimulation parameters. Only a few recent studies have started to systematically characterize the relationship between stimulation parameters and the perceived quality of artificial touch [25], [39]. When future technology enables us to selectively activate specific neurons in somatosensory cortex, likely creating more natural-feeling percepts, it would be interesting to repeat these tests.

Contrary to several previous studies [10], [13], [40], though similar to one [35], we found that both tactile stimuli were perceived slower than visual stimuli. This discrepancy may be caused by differences in task requirements (i.e. simple reaction time tests vs. acting on a stimulus), but many other factors influence reaction time [41]. The differences in reaction time between vision and touch are on the order of milliseconds and may be overshadowed by these factors given the small sample size of this study.

### Overlapping vibration and ICMS can be simultaneously perceived

To our knowledge, this is the first study that evaluated the perception of vibration and ICMS delivered such that they simultaneously activated overlapping populations of cortical neurons (by delivering vibration at the location of the projected field of ICMS, see Supplemental Figure 6). Though the participant performed well above chance in the modality discrimination task, the trials in which both vibration and ICMS were presented at the same time were still challenging (accuracy = 69%), likely due in part to vibration and ICMS engaging overlapping circuity in somatosensory cortex. Additionally, this discrimination difficulty may also result from vibration having a dominating effect over ICMS: when the participant misclassified stimuli in the simultaneous or -200 ms multiplexed conditions (vibration preceded ICMS), he reported feeling vibration only. When the two stimuli were temporally offset, identification accuracy improved; however, it is unclear if this improvement is the result of a longer total stimulation duration (i.e., 400 ms) making it easier to identify the two conditions or not. Further experiments would be needed to better quantify this effect. The participant’s accuracy was also significantly different between the two temporally offset multiplexed conditions (two-tailed two-sample binomial test, p=0.03); performance was higher when ICMS preceded vibration. Therefore, it appears that ICMS detectability was not completely overwhelmed by vibration, but it was substantially impaired.

### Disparity between reaction time and TOJ results

In contrast to the reaction time results, results from the TOJ task suggested that mechanically- and electrically-evoked sensations emerge with approximately the same latency. The participant’s difficulty discriminating between stimulus modalities may have hindered his performance on the TOJ task. Again, this confusion likely stems from the fact that the two stimuli engage overlapping neural circuitry in somatosensory cortex.

Note, however, that discrepancies between reaction time and TOJ tasks have been previously observed [42][43] and may reflect differences in task demands. Specifically, reaction time judgments are speeded whereas TOJ judgments are not. Accordingly, the nervous system may have time to resolve minor differences in sensory latency by the time the participant has to make a TOJ judgment. Another possibility is that the participant was biased towards reporting ICMS first because the evoked tactile percept did not feel natural, which required greater concentration to detect and interpret. This type of selective attentional bias can reduce detection time in a TOJ task [44]–[46].

### Limitations

Although we used previously validated experimental techniques, this study had a number of limitations. Our findings could become more generalizable if they were repeated in a larger sample size. Finally, though we matched the intensity between vibration and ICMS pairings, we did not match the intensities *across* pairings, which likely accounts for some of the cross-site variation (Supplemental Figures 3-5). It is also possible that because of the participant’s spinal cord injury, he had sensory deficits in his hands that could not be detected via clinical reports or personal awareness, which could have impacted the detectability of vibratory stimuli and caused differences in results across tested hand sites. Additionally, variations in stimulus duration in the modality discrimination task may have influenced the results. When vibration and ICMS were offset by 200 ms, the total trial length was 400 ms. It is possible that the participant noticed when stimulation duration was longer and subsequently reported feeling both stimuli, which may have unfairly increased his accuracy in that condition (which was 83% when vibration occurred first and 91% when ICMS occurred first). An interesting possibility for future work would be to systemically characterize the effect of other ICMS parameters, such as pulse frequency and pulse train length, on the temporal perception of ICMS.

### Implications for brain-computer interfaces

Somatosensory feedback improves performance on dexterous motor tasks [47], [48] while minimizing reliance on visual feedback [49]. Touch feedback provided via ICMS has recently been demonstrated to improve reach-to-grasp task completion times for a person with spinal cord injury [5]. Despite our observation that the perception of ICMS is slower than vibration, prior studies have demonstrated that ICMS perception can be accelerated by increasing stimulating current across at least four electrodes [10]. Future studies should be performed to evaluate whether high-intensity ICMS is comfortable, functionally beneficial, and detectable over long periods of time in sensorimotor tasks, simulating the use case of brain-computer interfaces (BCIs) outside of a laboratory. Additionally, the nervous system maintains multisensory synchrony by correcting for short time lags [34]. Even though the reaction time to mechanical tactile stimulation is slower than vision, these differences are compensated for, so the short delays observed in this study may not be problematic.

As one of the first studies in humans with simultaneous and overlapping touch stimuli delivered peripherally and to the brain, this study also has an impact beyond the scope of sensorimotor rehabilitation. A more precise understanding of how brain stimulation and natural touch interact will be critical for designing sensorimotor BCIs for able-bodied users in augmented reality or gaming applications [50], [51].

**Table 1:** Temporal order judgment task numerical results. The mean ± standard deviation of the point of subjective simultaneity (PSS) and just noticeable difference (JND) was calculated from the bootstrapped values. Positive PSS values indicate that ICMS needed to occur before vibration in order to be perceived as maximally simultaneous, and negative values indicate that vibration needed to occur first. After Bonferroni correction, none of the PSS values were statistically significantly different from zero. RR = right ring finger, RR = right thumb, LT = left thumb.

|  | **Spatially offset**<br>**vibration and**<br>**ICMS** | **Co-located**<br>**vibration and**<br>**ICMS** |
| --- | --- | --- |
| **PSS** |  |  |
| Vibrate RR | -12 ± 25 ms | -54 ± 24 ms |
| Vibrate RT | -57 ± 29 ms | +50 ± 34 ms |
| Vibrate LT | -50 ± 26 ms | +13 ± 34 ms |
| **JND** |  |  |
| Vibrate RR | 155 ± 25 ms | 133 ± 23 ms |
| Vibrate RT | 176 ± 28 ms | 230 ± 40 ms |
| Vibrate LT | 152 ± 27 ms | 216 ± 37 ms |

## Data Availability

All data produced in the present study are available upon reasonable request to the authors.

## Acknowledgements

The authors thank Pawel Kudela, Brock Wester, Manuel Anaya, and the research participant. This work was made possible, in part, through financial support from Defense Advanced Research Projects Agency (DARPA) under the Neurally Enhanced Operations program (contract number HR001120C0120) and the Johns Hopkins University Applied Physics Laboratory. In addition, this work was supported with the resources and use of facilities at the Johns Hopkins Hospital.

## Disclaimer

The views, opinions and/or findings expressed are those of the author and should not be interpreted as representing the official views or policies of the Department of Defense or the U.S. Government.

---

**Supplemental Figure 1:**
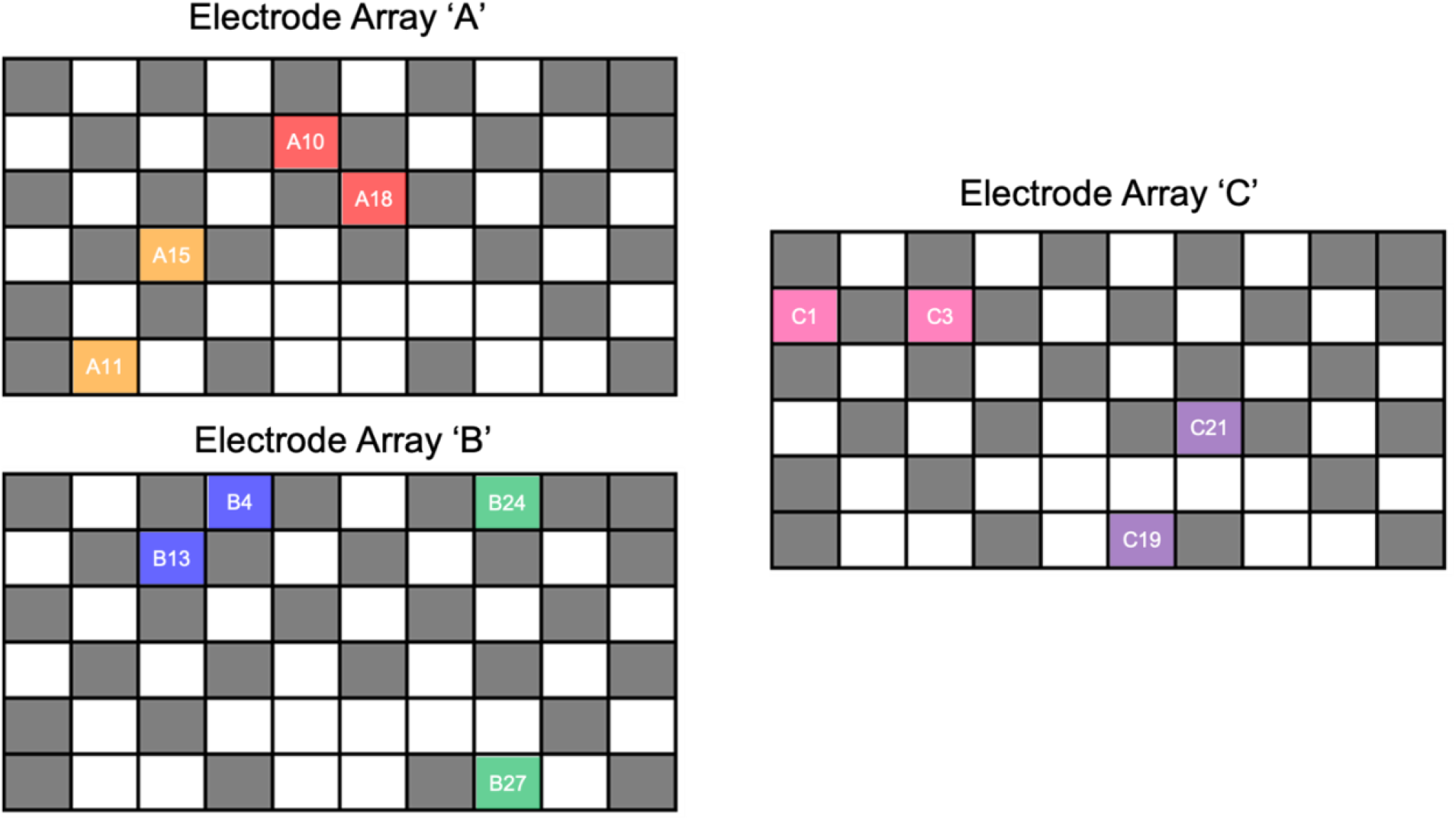
The BCI participant was implanted with two 6×10 electrode arrays in his left somatosensory cortex (arrays A and B) and one array in his right somatosensory cortex (array C). Each array contained 32 wired electrodes spaced 400 μm apart, and totaled 4 × 2.4 mm in size. Each electrode was 1.5 mm long and 80 µm in diameter. In the diagrams below, each square represents one electrode within the array; the gray squares depict disabled electrodes and white squares depict wired electrodes. The two electrode pairs shaded in red and pink were used in all three experimental tasks. The two pairs shaded in orange and green were used in the reaction time and modality discrimination tasks. The electrode pair shaded in blue was used in the TOJ and reaction time tests. The pair shaded in purple was used only in the TOJ task.

**Supplemental Figure 2:**
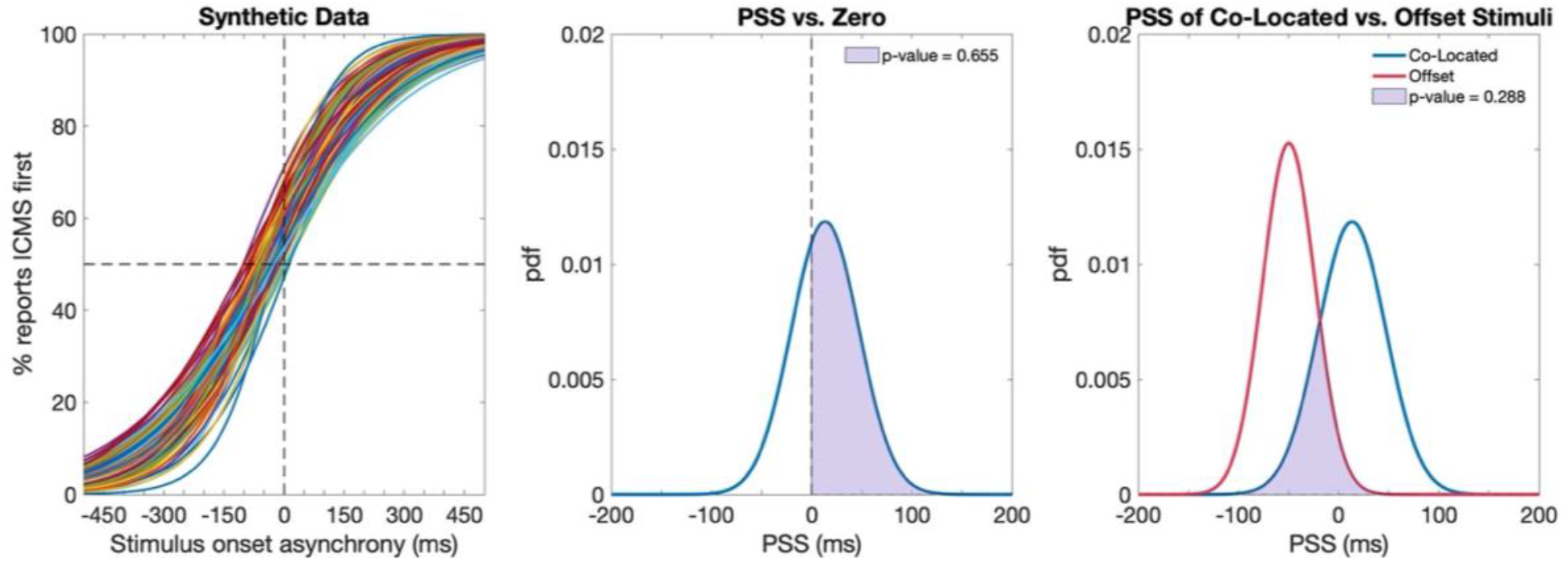
This figure lays out the analysis performed to analyze the results of the temporal order judgment task. **(a)** 1000 synthetic datasets were generated using a parametric bootstrapping method. The point of subjective simultaneity (PSS) and just noticeable difference (JND) were extracted from each synthetic dataset. **(b)** We fit a probability density function to the 1000 bootstrapped PSS values and calculated the proportion of samples that fell above zero, which was assumed to be the one-tailed p-value. A two-tailed test, to evaluate if PSS was significantly above or below zero, was performed by doubling the one-tailed p-value. **(c)** To compare the PSS and JND values between the spatially co-located vs. offset conditions, we calculated the proportion of overlap (taken to be the p-value) between the probability density functions of the two conditions.

**Supplemental Figure 3:**
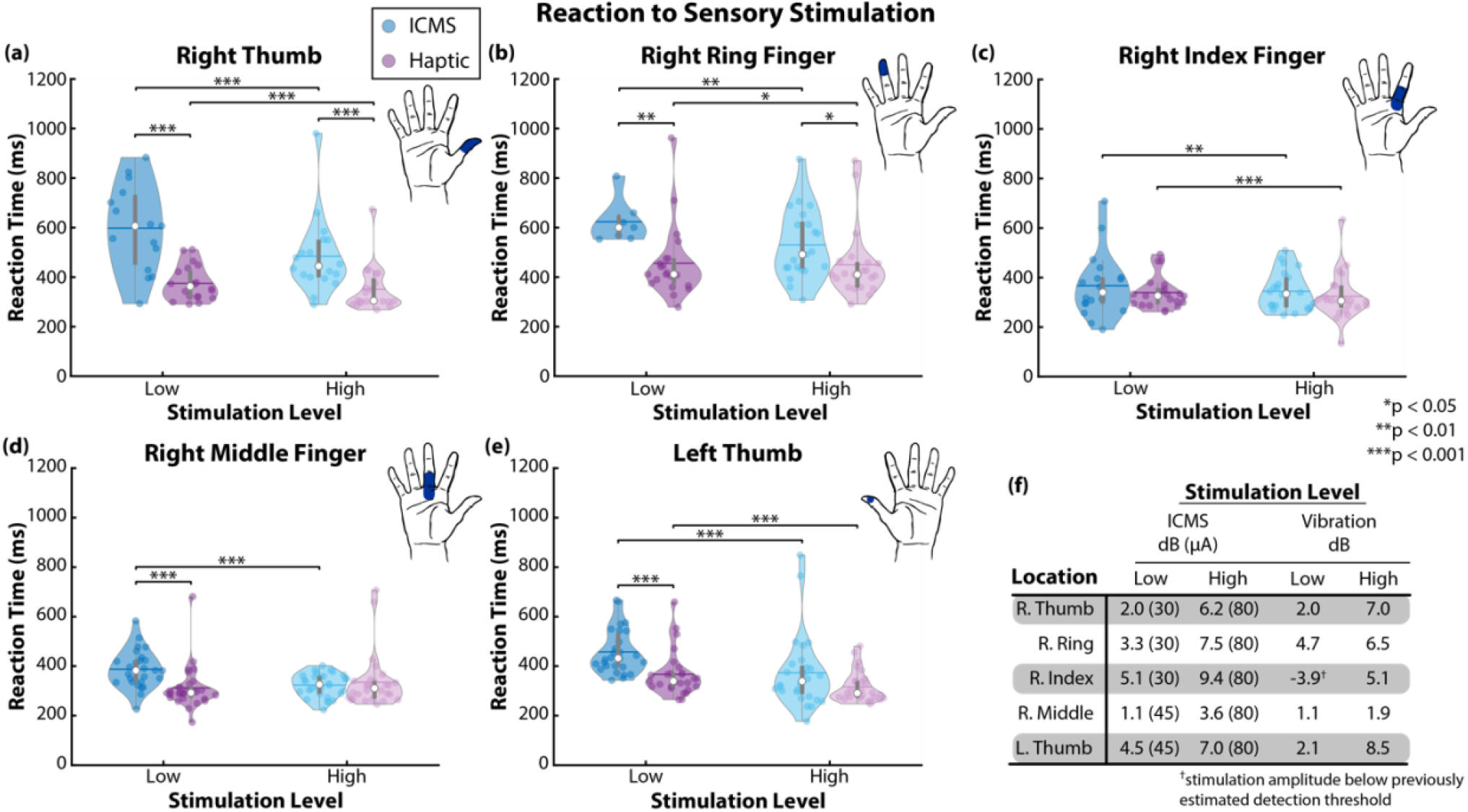
**(a-e)** Reaction time results for each site and perceived intensity level (“Low” and “High”) tested across both hands. In general, the participant’s reaction time to intracortical microstimulation (ICMS) was slower than his reaction to vibration at the same level of perceived intensity; however, this difference was more likely to be statistically significant at the lowest intensity level. Each point on the subplots represents one trial and the asterisks indicate statistically significant differences. **(f)** Vibration and ICMS levels used for each of the hand sites. Perceived intensity levels (“Low” and “High”) were matched between vibration and ICMS, but not across hand sites. Stimulation decibel levels are relative to previously estimated detection thresholds for that particular stimulation site.

**Supplemental Figure 4:**
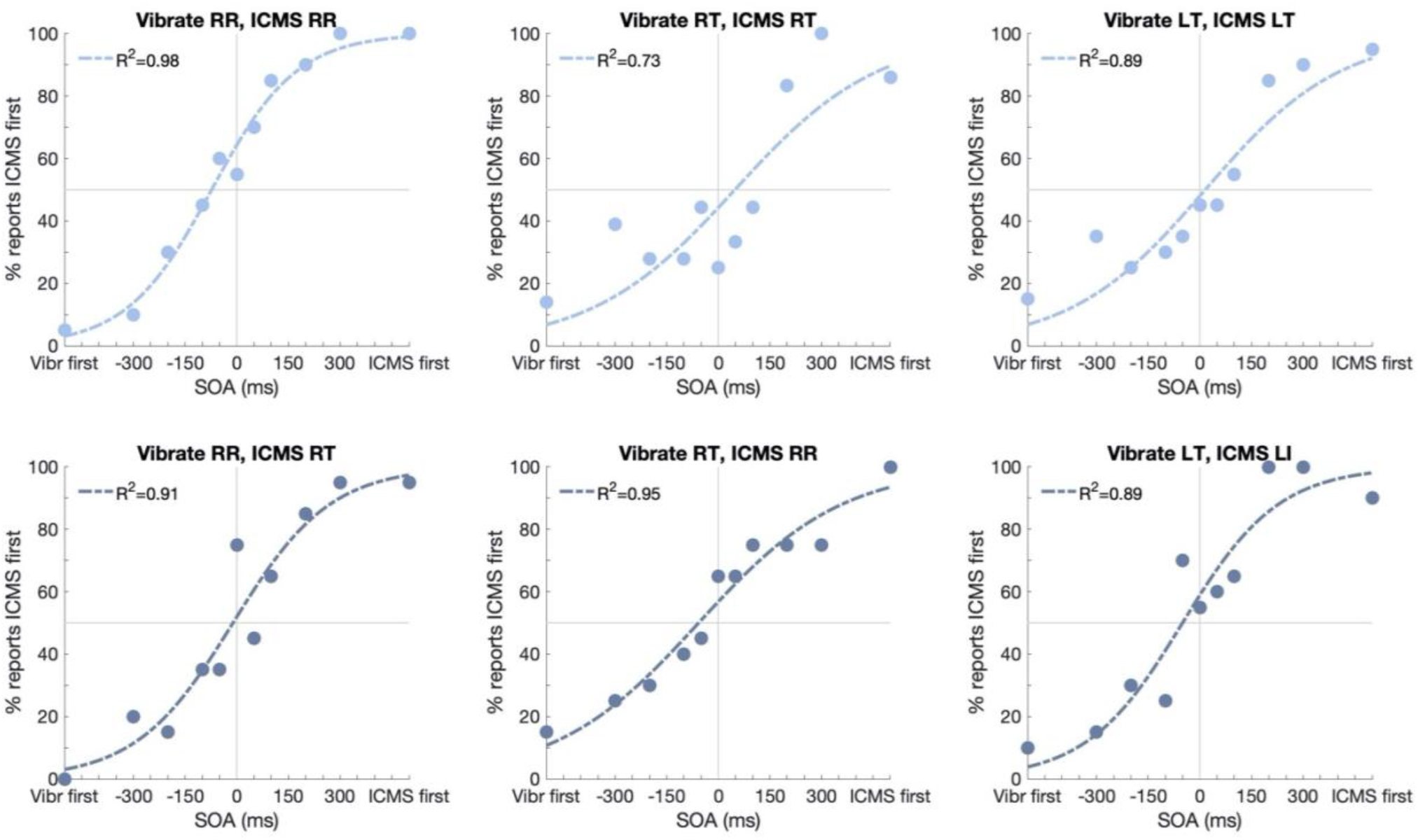
Temporal order judgment task raw data plus fitted curves using Equation 2. Positive stimulus onset asynchrony (SOA) values indicate that intracortical microstimulation (ICMS) occurred before vibration, and negative values indicate that vibration (‘Vibr’) occurred first. Each data point represents a percentage of 20 trials (except for the condition in which vibration and ICMS were both delivered to the right thumb, which involved 36 trials per SOA). Sigmoidal curves were fit to the raw data and goodness-of-fit R^2^ values are included on each plot. In the top row of plots, vibration and ICMS were delivered to overlapping regions of the hand. In the bottom row, vibration and ICMS were delivered to spatially offset regions of the hand. RR: right ring finger, RT: right thumb, LT: left thumb, LI: left index finger.

**Supplemental Figure 5:**
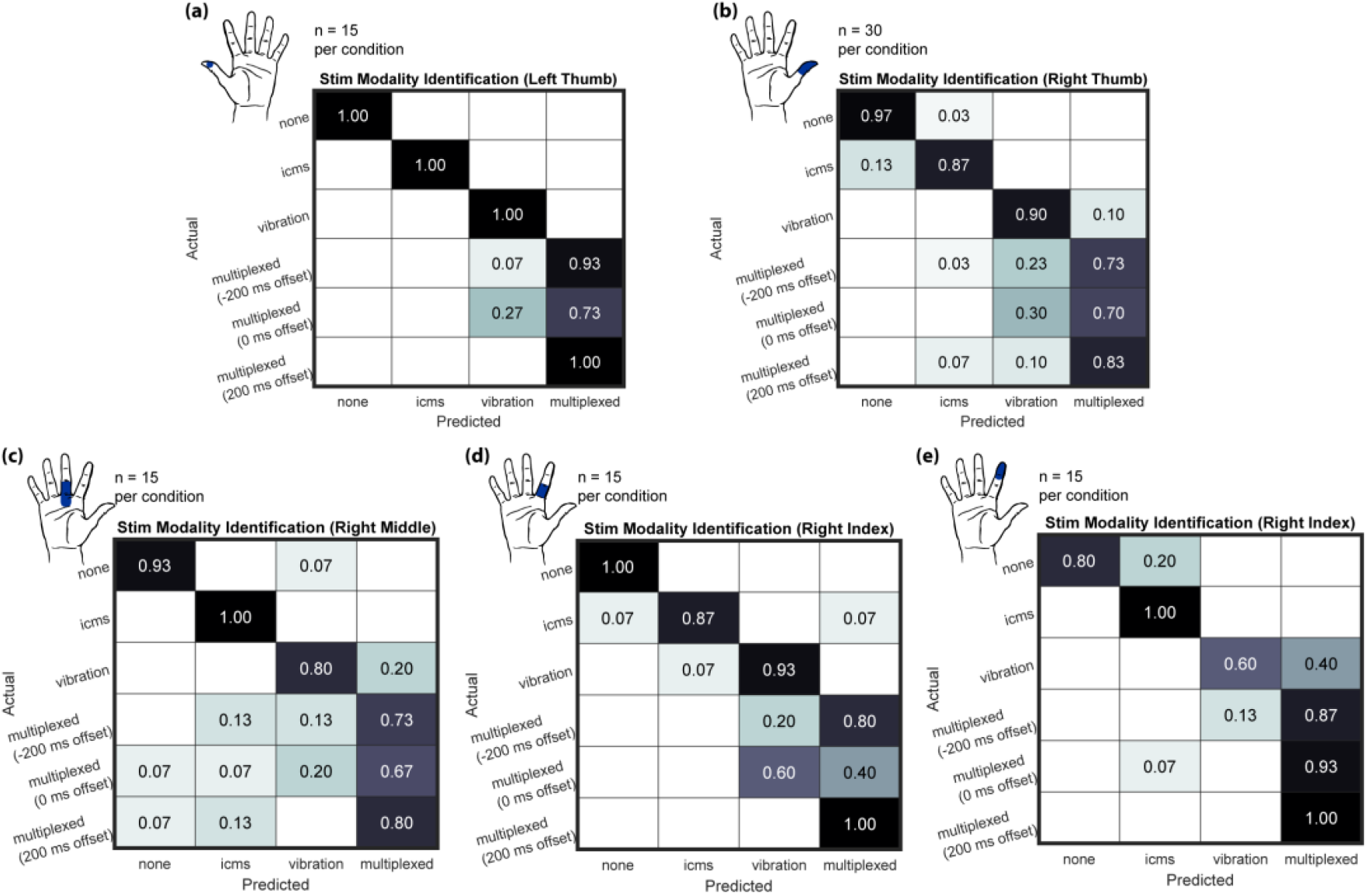
Modality discrimination task results across the four tested fingers: **(a)** left thumb, **(b)** right thumb, **(c)** base of the right middle finger, and the **(d)** base and **(e)** fingertip of the right index finger. It should be noted that the same ICMS electrode pair was used to activate both the **(d)** base and **(e)** fingertip of the right index finger. Although the same electrode pair was used on different days, the perceived focal point of activation in the right index finger differed slightly (i.e., base vs fingertip). The vibrating motor was placed accordingly to maximally overlap with the perceived ICMS sensation on each day. During the “multiplexed” condition, both vibration and ICMS were delivered. A negative temporal offset during the multiplexed condition signifies that vibration preceded ICMS and a positive offset indicates that ICMS preceded vibration. In all cases, the participant was able to correctly identify trials with only ICMS and only vibration at least 87% and 80% of the time, respectively. In general, the participant’s ability to accurately identify both stimuli modalities was lower compared to identifying a single stimulus modality, but performance ranged from as low as 40% (for simultaneous vibration and ICMS on the base of the right index finger) up to 100% for trials in which ICMS preceded vibration on the **(a)** left thumb and **(d)** right index finger.

**Supplemental Figure 6:**
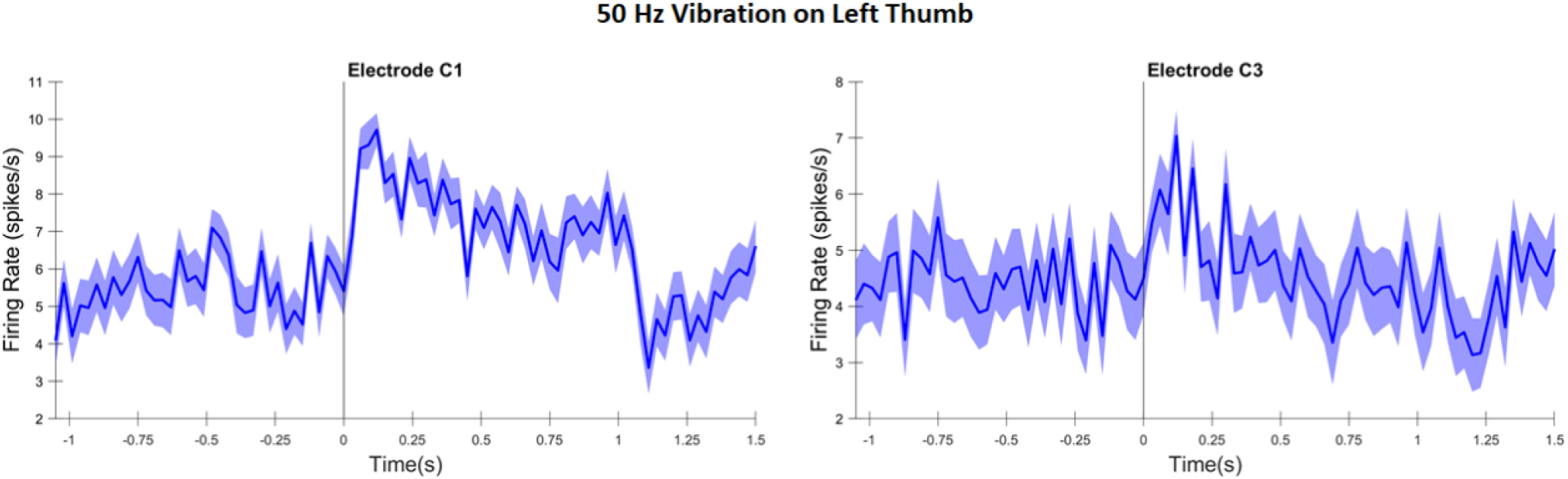
Firing rates recorded through **(a)** microelectrode C1 and **(b)** microelectrode C3 in the right hemisphere of the somatosensory cortex while a 50 Hz vibration was delivered to the participant’s left thumb. Vibration was delivered to the center of the distal phalanx (fingertip) using a custom-made Chubbuck vibrotactile stimulation device [52]. Vibration was delivered for 1 s for a total of 30 repetitions. Thresholded firing rates were recorded from both electrodes and segmented for a period of -1 s to +1.5 s relative to vibration onset. Action potentials were counted in 30 ms bins and then divided by 0.03 s to get the firing rates. The average firing rate across all repetitions is represented by the thick blue line and the standard error is represented by the blue shaded region.

**Supplemental Table 1:** The experimental conditions for all three tasks. The reaction time task and some conditions of the modality discrimination task presented only one stimulus per trial; therefore, we wrote “N/A” in the “Stimulus B” column. I_high_ and I_low_ signify that the test was performed once with a high-intensity stimulus and once with a low-intensity stimulus.

| Experimental task | Stimulus A | Stimulus B |
| --- | --- | --- |
| Reaction time | Vibration of right thumb ($I_{high}$ and $I_{low}$) | N/A |
| | Vibration of right ring finger ($I_{high}$ and $I_{low}$) | |
| | Vibration of right index finger ($I_{high}$ and $I_{low}$) | |
| | Vibration of right middle finger ($I_{high}$ and $I_{low}$) | |
| | Vibration of left thumb ($I_{high}$ and $I_{low}$) | |
| | ICMS of right thumb ($I_{high}$ and $I_{low}$) | |
| | ICMS of right ring finger ($I_{high}$ and $I_{low}$) | |
| | ICMS of right index finger ($I_{high}$ and $I_{low}$) | |
| | ICMS of right middle finger ($I_{high}$ and $I_{low}$) | |
| | ICMS of left thumb ($I_{high}$ and $I_{low}$) | |
| Temporal order judgement | Vibration of right ring finger | ICMS of right ring finger |
|  | Vibration of right ring finger | ICMS of right thumb |
|  | Vibration of right thumb | ICMS of right thumb |
|  | Vibration of right thumb | ICMS of right ring finger |
|  | Vibration of left thumb | ICMS of left thumb |
|  | Vibration of left thumb | ICMS of left index finger |
| Modality discrimination | Vibration of left thumb | N/A |
|  | Vibration of right thumb |  |
|  | Vibration of right middle finger |  |
|  | Vibration of right index finger |  |
|  | Vibration of left thumb | ICMS of left thumb |
|  | Vibration of right thumb | ICMS of right thumb |
|  | Vibration of right middle finger | ICMS of right middle finger |
|  | Vibration of right index finger | ICMS of right index finger |
|  | ICMS of left thumb | N/A |
|  | ICMS of right thumb |  |
|  | ICMS of right middle finger |  |
|  | ICMS of right index finger |  |
|  | None (control condition) |  |

